# Predicting the COVID-19 positive cases in India with concern to Lockdown by using Mathematical and Machine Learning based Models

**DOI:** 10.1101/2020.05.16.20104133

**Authors:** Ajit Kumar Pasayat, Satya Narayan Pati, Aashirbad Maharana

## Abstract

In this study, we analyze the number of infected positive cases of COVID-19 outbreak with concern to lockdown in India in the time window of February 11th 2020 to Jun 30th 2020. The first case in India was reported in Kerala on January 30th 2020. To break the chain of spreading, Government announced a nationwide lockdown on March 24th 2020, which is increased two times. The Ongoing lockdown 3.0 is over on May 18th, 2020. We derived how the lockdown relaxation is going to impact on containment of the outbreak. Here the Exponential Growth Model has been used to derive the epidemic curve based on the data collected from February 11th 2020, to May 11th 2020, and the Machine Learning based Linear Regression model that gives the epidemic curve to predict the cases with the continuous flow of the lockdown. We estimate that if the lockdown is continuing with more relaxation, then the estimated infected cases reach up to 1.16 crores by June 30th 2020, and the lockdown would persist with current restriction, then the expected predicted infected cases are 5.69 lacs. The Exponential Growth Model and the Linear Regression Model are advantageous to predict the number of affected cases of COVID-19. These models can be used for forecasting in long term intervals. It shows from our result that lockdown with certain restriction has a vital role in preventing the spreading of this epidemic in this current situation.

## Introduction

The novel coronavirus puts the world to go through its distress stage. The World Health Organisation (WHO) is declared this situation as a global public health emergency. In the last two decades, more than 10,000 people in additive order are affected by the contagious β-coronavirus, which is known as Severe Acute Respiratory Syndrome Coronavirus (SARS-COV) and the Middle East Respiratory Syndrome Coronavirus (MERS-COV) [1]. According to the Centre for Disease Control and Prevention (CDC), the recent coronavirus has some Correspondence with SARS-COV and MERS-COV [2]. The first case was reported as unidentified pneumonia in the People’s Republic of China (PRC) in late December 2019 [2], which later detected as coronavirus. WHO was later on March 11^th^ 2020, framed it as Pandemic[1]. Then as time flows, the number of coronavirus victims spread exponentially. WHO named this novel coronavirus as “COVID-19” on February 11^th^ 2020 [3].

The initial symptoms are fever, cough, and shortness of breath. Apart from this, it may include pneumonia and acute respiratory distress syndromes. Now, in India, a lot of positive cases are found which do not have any symptoms [4]. This Contagious virus spread while the persons are closer to each other. It takes the medium of small droplets produced by coughing, sneezing, or talking [5]. And also, it can spread via passive interconnection with the droplets. This epidemic may be causing respiratory illness, throat infection, and affect the lungs, which may be the reason of death.

Due to smooth transmission, it is rapidly spreading overworld. 212 Countries and Territories around the world are under threat of this diesis [6]. There is a total of 4,362,016 infected cases since May 12^th^ 2020, worldwide and 293,304 deceases cases, along with 1,613,326 recovered cases, are reported [6]. It is difficult to identify this disease at the preliminary stage, so the actual number of cases would be much higher [7]. According to the report, twelve to eighteen months are required to make vaccines of COVID-19 [8]. In this contingency, the major preventive guidelines are delivered by WHO. The spreading of the virus to enter into our body can be stopped by regular hand wash with alcohol-based hand rub or wash with soap and water, maintain at least 1 meter (3 feet) of social-distance, avoid touching eyes, nose, face [9]. The essential measures should be taken by Government to perpetrate social-distancing. Lockdown plays a crucial role in breaking the rapid spreading of this virus. It provides restrictions on goods transportations, closing or minimizing the travel connections from different parts of the country, closing the public place, tourist spots, vending zones, cultural activities plays a crucial role in disrupting the spreading of this deadly disease. The schools, colleges, universities are closed in most of the countries. Though all the business activities are shut down, the whole world faces massive loose in the economy.

India is the 2nd largest populated country in the world. It is having 17% of the world’s total population, and it is also among very high population-dense countries. India is now affected by COVLD–19. By studying the rate of spread of this virus in other countries, it shows that till now India has taken the appropriate prevention steps like lockdown in 3 stages, for which the rate of spread of this novel coronavirus is less.

The first positive case of COVID-19 in Kerala, India, was reported on January 30 2020, which had an originating history of China [10]. According to the Health Ministry of India, it is in 2nd to 3rd stage of this Pandemic [11]. By May 12^th^ 2020, there is a total of 75,048 corona positive cases in India, out of which 2,440 fatal cases and 24,900 the recovered cases are filed [12]. It produces the Case Fatality Ratio (CFR) up to 3.3 percent on April 18^th^ 2020 [13]. Now in most part of India community transmission is not seen yet. The number of hospitals has to be sufficient to deal with any disaster. According to the report, there is less than 1 hospital bed available per 1000 people in India [8]. As reported early, there is still no vaccine available for COVID-19, so social-distancing and lockdown are the vital strategies that have been taken. The number of tests should be improved to find out the positive COVID-19 cases which help to break the chain. These steps could help to stop the spreading of COVID-19 efficiently.

In India, to control the spreading of this deadly virus, Government announced 3 consecutive nationwide lockdowns of 40 days on March 24^th^ 2020 to May 18^th^ 2020[14]. It has a crucial impact on breaking the chain, which causes the slowdown of the spreading of this contagious virus. This lockdown also gives precious time to make temporary medical camps, COIVD–19 special wards in the hospital, increase the number of medical facilities, produce a sufficient amount of testing kits, and Personal Protection Equipment (PPE) for health care workers.

The Government recognize different part of the country into red, orange and green zones with respect to the effectiveness of this virus. In the current lockdown phase, 3.0, the Government of India has provided the railway facility to the migrants who are staying in different part of the country. The migrants travel from one part of Country to other part without being test of COVID-19 which causes the virus to spread very rapidly. Also, the railway service is available to the general citizens in some regions. Recently the Government plans to increase the lockdown with more relaxations which may be a great thread to the spread of this virus.

In this paper, we predict and compare the positive cases in concern to relaxation in lockdown. For this we proposed a mathematical based Exponential Growth Model and a Machine Learning based Linear Regression model to calculate the growth rate of the COVID-19 positive case in India with concern to lockdown.

## Methods

### Data Source

We downloaded the publicly available dataset from Humanitarian [15]. It contains the day-wise number of cases, deaths, recovered of all the corona affected countries form which Indian data is extracted for our study. After processing, we avail the data publicly at data. World [16].

### Models

In this paper, we are using two different models for predicting the positive cases till June 30^th^ 2020, with concern to lockdown. The two models are Mathematical Exponential Growth Model, and Machine Learning based Linear Regression Model. The Exponential Growth Model is used to predict the number of positive cases if the lockdown is not continuing strictly after May 18^th^ 2020, and the Linear Regression Model is used to predict the number of positive cases with concern to lockdown continue.

### Exponential Growth Model

Exponential Growth is a mathematical function which tells us the number of cases at a specific moment in time. According to the researcher, the first period of epidemic follows exponential growth. This is the reason to use this mathematical model for the Coronavirus outbreak [17].

In this model, the quantity increases over a period. The quantity is proportional to the derivative of the quantity with respect to time. A quantity undergoing exponential growth is the exponential function of time. That means the time variable is the exponent.

We observed the growth of COVID-19 outside China is following exponential growth by plotting the number of cases against time.

The model can be described as:

ΔP_d_ = Number of new cases on a given day d.

q = Average number of people someone infected is exposed to each day.

r = Probability of each exposure becoming an infection.

P_d_ = Number of total cases on a given day d.

The number of cases on d+1 day is formulated as

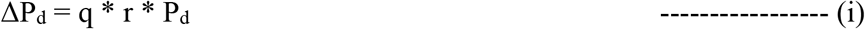

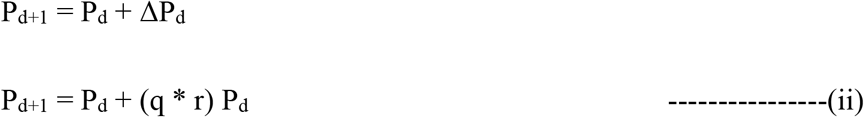

Then the number of cases on d+2 day will be

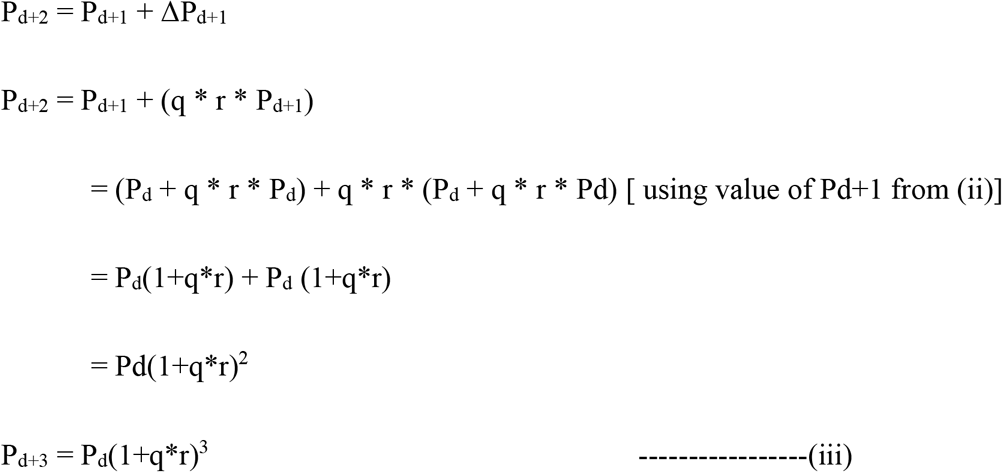

We can generalize the equation as,

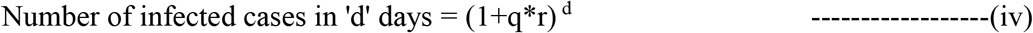

The general equation of an exponential model [18] is

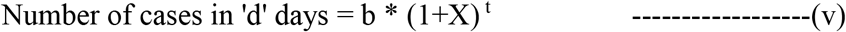

1+X = growth factor

b = number of cases in first occurrence [=1 in India]

t = Time factor [here t = number of days]

Equation (5) is a general equation of exponential growth.

By comparing equation (iv) to (v) we can get that X = q * r.

### Estimation of Model Parameters of Exponential Growth Model

As there is no mass testing in India, so taking the aggregate data may be problematic. To address the problem, we resample the data to the Inter Quantile Range [25% to 75%]. In the resampled data, we found the mean growth factor 1.106. After iterating with an error range of 5%, we got the best fit line. In the best fit line, the mean growth factor is 1.115.

### Result Analysis

The equation (v) gives the best fit line with a mean growth factor of 1.115. We calculated the r2 score [17] and got a result of 90.78. Assuming there is no strict lockdown after May 18^th^ 2020, in the best-case scenario, the growth factor remains the same as of now shows in Figure-1. If, even after the lockdown, the Government enforces social distancing, isolation of containment zones, and a ban on public gathering and other preventive measures, then the growth factor may reduce. Still, lockdown with more relaxation, we may face around 1.16 crores cases in worst scenario. Else, if the exposure of infected people increases, the growth factor may increase and the situation is out of control.

**Figure 1.**
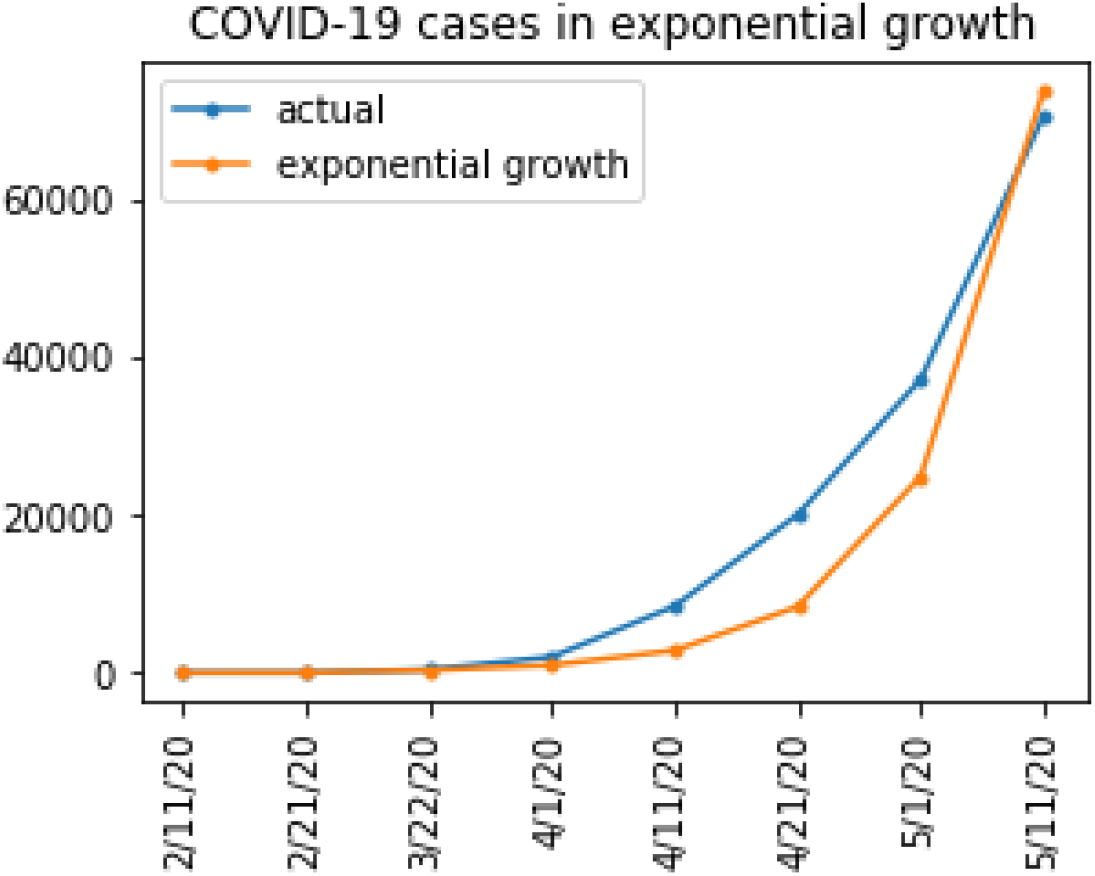
The Number of positive cases according to Exponential Growth Model represented with orange line and the Actual Number of Positive cases represented with blue line. Here Y axis is represented for Number of Positive cases and X axis represented the Date.

Though there is no real exponential model that works in epidemics, after a certain period, it has to be flattened. In the worst case, all the possible number of people infected. After that, the curve has to be flattened. So, in the epidemic, the exponential growth model follows the path of a logistic curve. It increases until the inflation point. At the inflation point, the growth rate becomes 1, and the derivative becomes 0. After the inflation point, the growth factor may be less than 0, and the derivative is negative. And when the growth factor tends to 0, the line flattens. The total number of cases is double the number of cases at an inflection point.

### Linear Regression Model

In Linear Regression, the relationship is in between one dependent variable and one or more independent variables [19]. Here we are using an independent variable as the number of days, and the dependent variable is the infected positive cases.

By applying sklearn.preprocessing.PolynomialFeatures [19] we convert a single independent feature to a 4-degree polynomial feature to the regression curve with the hypothesis. Then we divide our data into train and test sets with the ratio 8:2. The model is trained with training data and validated against test data by using the following equation generated by the regression model [19].

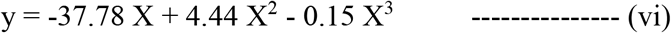

### Estimation of Model Parameters of Linear Regression Model

Here basically 2 parameters are used:

y = number of estimated cases,

X = number of days.

## Result Analysis

The linear regression model follows the trend in data. The model produces an accuracy (r2 score) of about 99.94% in training data and 99.89% in testing data. This model trained to predict the worst possible number of positive cases if there is no strict lockdown after May 18^th^ 2020 as shown in Figure-2. We estimated using this model and found that we may face about 5.69 lacs number of positive cases in the best-case till June 30^th^ 2020. Assuming the lockdown would be extended, the Government would be able to give less relaxation and focus on more social distance, conduct more tests find possible positive cases, isolating and quarantining the suspected and taking other preventive measures. So, the situation could be under control, and the growth might be flattened.

**Figure 2.**
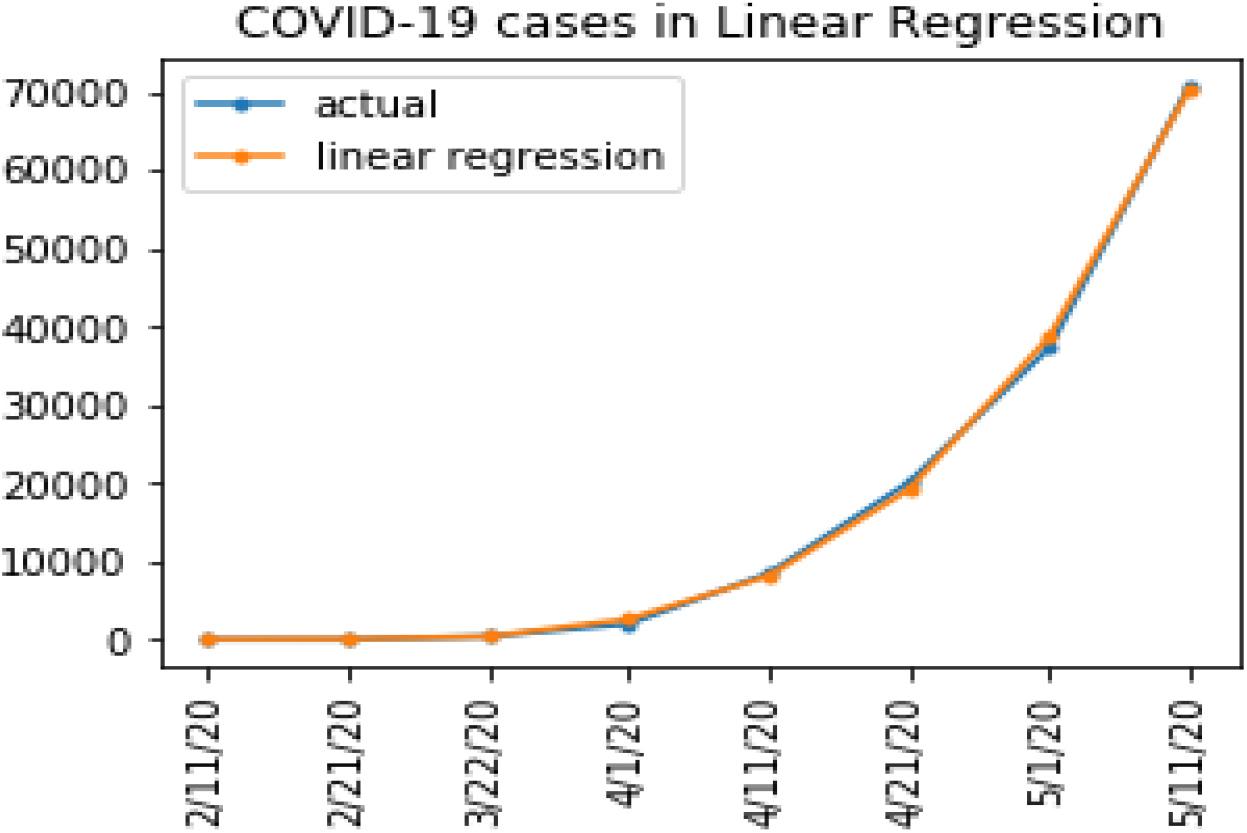
The Number of positive cases according to Linear Regression represented with orange line and the Actual Number of Positive cases represented with blue line. Here Y axis is represented for Number of Positive cases and X axis represented the Date.

## Performance

The efficiency of an estimator can be measured with its performance. We take r2_score [19] as a performance measure for both the models. The variation occurs in a linear model explain as percentage of the dependent variable which measure as r2_score. The model that does not make any of the variations in the response variable around its mean then it is represented as 0% and if it makes all of the variations, then it is represented as 100%. Our model gives the accuracy as shown in Figure 4:

## Discussion

As the government takes control measures, we observe a linear growth in case if lockdown with minimize relaxation will extend. And also, we observe that the Linear Regression growth model is more specific than the Exponential Growth Model. Exponential Growth Model fits if there is lockdown with relaxation.

During the process of data, there were some challenges, as there is no mass testing in India, so using aggregate data may be problematic for the prediction, which also occurred in the exponential prediction model. Predicting by taking days as only dependent vectors is also questionable, but we investigate the trend in our data. To overcome the first challenge, we resample the data by taking the Interquartile Range (IQR range) [25% to 75%] and find the growth factor. To address the second problem, we create polynomial features with the help of sklearn.preprocessing.PolynomialFeatures. The 3rd problem is predicting with 90 samples not suitable for machine learning. However, we test with 20% of the dataset 21 samples and achieve 99.89% accuracy in regression and r2 score of 90.78% in the exponential model as shown in Figure-4. Due to the shortage of data, there might be a problem of overfitting, but we achieve train test accuracy and test set accuracy nearly the same.

In this paper, we only predict the number of infected positive cases in the future, but don’t predict the recovery and death cases. The recovery and death of positive cases depend on various factors like underlying diseases in a positive case, age of positive cases, and also the recovery of an infected person also depends on the average age group infected and the immunity system. So we only limited our prediction of the number of infected people.

However, the growth rate might have some variance, but outside China, the COVID-19 is growing exponentially. That’s why we implement the exponential model for India. We estimate that if the lockdown is more relaxed, then the spreading of this virus is in exponential format and reaches to 1.16crores and if strict lockdown followed the number of cases restricted to 5.69 lacs. The predict value shows in figure-3.

**Figure 3.**
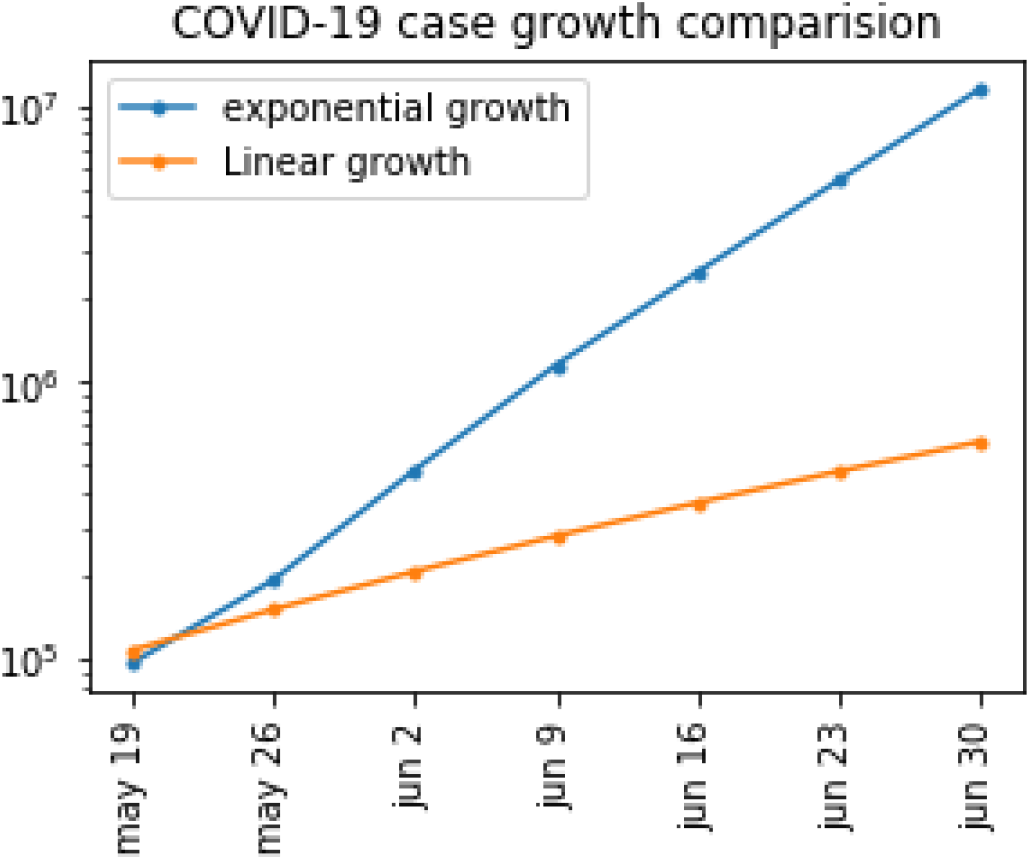
This figure is the Prediction of the number of positive cases by the two proposed models from May 19^th^ 2020 to Jun 30^th^ 2020. Here Exponential Growth Model (blue) and Linear Regression Growth Model(orange) lines show the huge gap, which is generated if the lockdown is more relaxed. X axis represents the Number of Positive cases and Y axis represents the Date.

**Figure 4.**
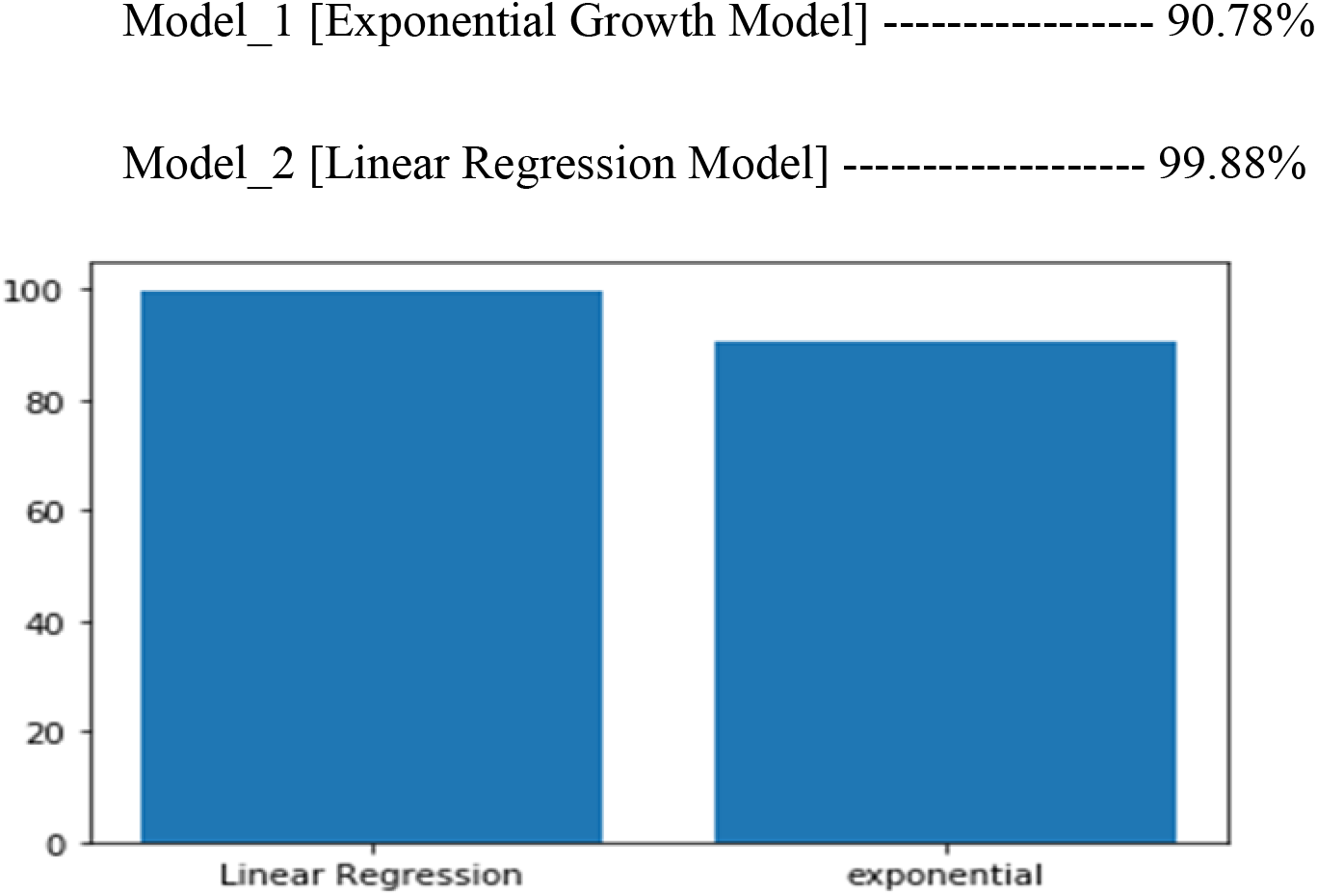
Comparision the performance Exponential Growth Model and Linear Regression Model.

## Conclusion

In this paper, we have implemented the proposed Exponential Growth Model, and a Machine Learning based Linear Regression Model. With these models, we show the variation of the growth rate of positive cases in India concerning the strictness of lockdown. Here the Exponential Growth Model exposes the contagious nature of the disease. On the other hand, the Machine Learning model estimates the cases based on the lockdown condition. In different states of India, a lot of hotspots are created where the positive cases are rapidly increasing. In this condition, if the lockdown is withdrawn or more relaxed, then it creates a disaster situation. This also leads to bothering in other precautions as social-distancing, self-quarantine. From the analysis, we conclude that there is a significant gap in the positive cases on the condition of lockdown. Earlier, we have cleared that the lockdown is the higher impactable precaution measure that has been taken in different countries of the world. The lockdown not only maintains social-distancing but also gives some safe time for the agencies to increase the preventive measure tools, kits, and make more tests. In this crucial situation, these models give useful insight, which may help to take appropriate steps to save the people from spreading the deadly disease.

## Data Availability

The data are collected from publicly available domains
on the internet.

https://data.humdata.org/dataset/novel-coronavirus-2019-ncov-cases

https://data.world/gabbarsingh/covid19india

